# Exploring the Causal Relationship Between Body Mass Index and Kidney Function Using Tissue-Partitioned Mendelian Randomization

**DOI:** 10.1101/2025.08.19.25333885

**Authors:** Artemis Briasouli, Genevieve Leyden, Aristomo Andries, Stein Ivar Hallan, Tom G. Richardson, Bjørn Olav Åsvold, Humaira Rasheed, Ben Michael Brumpton

## Abstract

**Background:** Chronic kidney disease (CKD) represents a leading non-communicable disease, significantly contributing to global morbidity and mortality. Mendelian randomization studies (MR) have been integral in providing robust evidence that increased body mass index (BMI) has a causal impact on CKD. However, dissecting which specific mechanisms are primarily responsible for disease development remains challenging.

**Objective:** To explore whether the effects of BMI to kidney function are driven primarily by brain-or adipose-tissue derived gene expression using tissue-partitioned Mendelian randomization (MR).

**Methods:** We employ two-sample univariable and multivariable MR methodology that segregates genetic variants associated with BMI based on colocalization with gene expression in either brain or subcutaneous adipose tissue. We utilize sets of adipose and brain expression quantitative trait loci (eQTLs) that demonstrated colocalization with BMI (86 and 140 loci respectively). We also use GWAS summary statistics of creatinine and cystatin C based eGFR (eGFRcrea and eGFRcys; N=460,826), blood urea nitrogen (BUN; N = 852,678), eGFR decline (N= 34,874 cases) and CKD (defined as eGFRcrea <60=ml=min^−1^ per 1.73=m^2^; N= 41,395) of European ancestry.

**Results:** Univariable MR showed consistent positive associations between BMI and CKD (OR = 1.24, 95% CI: 1.2–1.3) and inverse associations with eGFRcys (beta = –0.05, 95% CI: –0.06 to –0.046). Both brain-and adipose-instrumented BMI showed similar effect sizes. However, in multivariable MR, neither brain-nor adipose-specific BMI variants showed clear independent effects on CKD (OR_adipose_= 1.24, 95% CI_adipose_= 0.87 to 1.65, OR_brain=_ 1.18, 95%CI_brain_ = 0.9 to 1.54) or other kidney function traits.

**Conclusions:** While genetically predicted BMI was associated with kidney function, our tissue-partitioned MR analysis found no strong evidence that brain or subcutaneous adipose tissue derived gene expression independently drive this relationship. This suggests overlapping or additive mechanisms through which BMI influences kidney function.

## Introduction

Obesity, defined by a body mass index (BMI) of ≥30 kg/m², is a major public health challenge globally, contributing to the development of numerous chronic diseases, including type 2 diabetes (T2D), coronary artery disease (CAD), and chronic kidney disease (CKD) [1].

While the causal relationship between obesity and CKD has been established through both traditional observational studies and Mendelian randomization (MR) studies [2,3,4], the underlying mechanisms are complex and remain incompletely understood. BMI, often used as a measure of adiposity, is a composite trait influenced by fat and lean mass, but also other factors such as bone density [5]. This heterogeneity complicates efforts to understand the causal pathways to disease.

Genetic studies have identified over 900 loci associated with BMI, highlighting its polygenic nature and significant genetic component [6]. Intriguingly, the functional roles of these genetic variants appear to vary by tissue type, with most BMI-associated loci showing high expression in neural tissue, suggesting a central role of the brain in appetite regulation and body composition [7]. Of note emerging evidence from recent studies highlights the involvement of multiple brain regions in modulating susceptibility to obesity [8,9,10], extending beyond the traditionally recognized role of the hypothalamus in appetite regulation, particularly in monogenic and syndromic forms of severe obesity. However, other findings point to adipose tissue as a critical mechanism particularly in the context of fat distribution and metabolic health. For example, genetic variants linked to higher subcutaneous fat storage capacity have been associated with a reduced risk of cardiometabolic diseases, underscoring the importance of fat depot-specific effects [11]. This suggests a metabolically healthy obesity phenotype where increased fat is not necessarily harmful due to favourable fat distribution [12].

Previous work has proposed that the considerable heterogeneity observed in BMI-related Mendelian randomization analyses [13] might be mitigated by partitioning BMI-associated variants according to their tissue-specific gene expression profiles, particularly in brain and adipose tissue [14]. This tissue-partitioning approach may help develop mechanistic understanding into BMI’s causal pathways towards disease risk [14]. Building on this concept and while the causal link between BMI and CKD has been supported by MR studies [15], the role of tissue-partitioned BMI, specifically brain-and adipose-mediated effects, in CKD development warrants further investigation. Therefore, the aim of this study is to investigate the potential causal effects of brain-and adipose tissue–partitioned BMI on CKD and related kidney function markers, including eGFR and eGFR decline, using Mendelian randomization, to elucidate tissue-specific mechanisms underlying the relationship between BMI and kidney health.

## Methods

### Study Design

We employed a two-sample MR study using both univariable and multivariable Mendelian randomization (MVMR), which estimates the independent effects of multiple exposures on an outcome by evaluating their effects simultaneously within a single model [16,17].

Univariable MR and MVMR aim to test causal effects of an exposure on an outcome. Specifically, MR estimates the total effect of the exposure on the outcome, while MVMR determines the direct effect of each individual exposure on the outcome accounting for each other exposure in the model. Conventional univariable MR can yield consistent results when the selected SNPs are valid instruments for the exposure [18]. MVMR is an extension to MR which facilitates the estimation of independent direct effects of multiple, potentially correlated exposures [16,19]. This is achieved by the estimation of each exposure on the outcome conditional on the other exposure included in the model. In this study, we applied MVMR to examine the tissue-specific effects of BMI by adipose and brain tissue, independently of one another, on each of the kidney outcomes under investigation.

We applied a novel MVMR approach from Leyden et al. [14,19], to explore whether the relationship between BMI and kidney outcomes is primarily driven by subcutaneous adipose or brain-tissue derived gene expression. Notably, the estimates derived from tissue-specific instruments should not be interpreted as direct causal effects in the traditional sense of Mendelian randomization analyses. Rather, this method was designed to disentangle the distinct contributions of genetic instruments reflecting different aspects of a single complex trait. In this study, BMI served as an exemplar, partitioned into adipose-and brain-tissue-derived gene expression pathways.

### Tissue-specific genetic instruments for BMI

We used genetic associations for BMI from a large-scale meta-analysis combining data from the Genetic Investigation of Anthropometric Traits (GIANT) consortium and the UK Biobank (N = 681,275) [6]. Independent BMI-associated SNPs were identified through linkage disequilibrium clumping (on the basis of p < 5 × 10^−8^). Based on a reference panel of 10,000 unrelated individuals (r2 < 0.01) of European ancestry from the UK Biobank, 915 independent variants were robustly associated with BMI (Supplementary Table ST2) [20].

We utilized tissue-partitioned colocalization results (Supplementary Table ST1), which examined the genetic overlap between BMI-associated variants and tissue-specific gene expression in brain and adipose tissues [14]. For brain, meta-analyzed brain eQTL data from Qi et al. [21] were used, which combined data from GTEx v7 [21], the CommonMind Consortium (CMC) [23], and ROSMAP (n = 1,194) [24]. For adipose tissue, meta-analyzed subcutaneous adipose eQTL data from the MuTHER study [25] and GTEx v8 (European ancestry, total n = 1,257) [26] were utilized. All eQTL data were mapped to the GRCh37/hg19 reference genome. Additional details on the sources and characteristics of these datasets are provided in Table 2. Briefly, Bayesian colocalization (coloc) was used to assess whether BMI genetic variants colocalized with gene expression of brain and adipose tissue based on a 200 kb cis-window, providing posterior probabilities of shared causal variants. Results with a posterior probability (PPA4) > 0.8 were considered evidence of colocalization. This yielded 140 brain-instrumented and 88 adipose-instrumented SNPs. Further details are provided in Supplementary Table ST1. Additional details for colocalization and tissue-specific gene expression meta-analysis can be found in Leyden *et al*. [14]

**Table 2.** Description of the GWAS data used in this study.

| Phenotype | Phenotype definition | Cohort | Sample size | Population |
| --- | --- | --- | --- | --- |
| Exposure |  |  |  |  |
| Brain cis-eQTL (Qi |  | Meta-analyzed gene | 1194 | European |
| *et al.*, 2022) |  | expression datasets derived from brain tissue using GTEx v6, ROSMAP, CMC |  |  |
| Adipose cis-eQTL (Leyden *et al.*, 2022) |  | Meta-analyzed gene expression datasets derived from subcutaneous adipose tissue using Muthur, GTEx v8 | 1257 | European |
| Outcome |  |  |  |  |
| Continuous kidney function (Stanzick *et al.*, 2021) | eGFRcrea<br>eGFRcys<br>BUN | Meta-analyzed GWASs of UKB and CKDgen | 460,826 | European |
|  |  |  | 1,004,040 |  |
|  |  |  | 852,678 |  |
| eGFR decline (Gorski *et al.*, 2022) | (eGFR at follow-up – eGFR at baseline) / years of follow-up | GWAS meta-analysis of 62 studies (adjusted for age, sex and diabetes-status) | 343,339 individuals | Mostly European (74%) |
| CKD (Pattaro *et al.*, 2016) | eGFRcrea<br><60 ml min<sup>-1</sup> per 1.73 m<sup>2</sup> | GWAS meta-analysis of 49 studies | 117,165 including 12,385 CKD cases | European |
| CKD (Wuttke *et al.*, 2019) | eGFR | GWAS meta-analysis CKDgen and MVP >1 million participants | 41,395 cases and 439,303 controls | European |
| BMI (Jengo *et al.*, 2018) |  | GWAS meta-analysis | 681,275 | European |

### CKD-related outcomes

Our kidney related GWAS instruments were based on kidney function expressed as continuous variables (eGFRcr, eGFRcys, and s-BUN), slope (eGFR change per year (ml/min/1.73m2 /year), and dichotomized variables (CKD defined as eGFR <60 ml/min/1.73m2). In these datasets, positive effect estimates indicate higher trait values with higher BUN, higher slope, and lower eGFR values indicating a worse prognosis. First, we used continuous cross-sectional data from Stanzick et al. of predominantly European ancestry from the UK Biobank and CKDGen [27]. eGFR was based on serum creatinine (N=852,678) or cystatin C (N=460,826), and in addition blood urea nitrogen was reported (BUN; N = 1,004,040), eGFR measures were log-transformed using the natural logarithm (ln). BUN values were first multiplied by 2.8, then natural log-transformed to obtain beta estimates. The units for eGFR and BUN were ml/min/1.73 m² and mg/dL, respectively. Second, we utilized GWAS summary statistics from Gorski et al. [28], which estimated the annual decline in kidney function based on creatinine-derived eGFR. This GWAS, conducted by the CKDGen Consortium [28], included 334,339 individuals, of whom 74% were of European ancestry.

Annual eGFR change was calculated using the formula:-(eGFR at follow-up – eGFR at baseline) / years of follow-up. Of note, annual decline in eGFR was adjusted for age, sex, and diabetes status. These summary statistics can be considered equivalent to those adjusted only for age and sex, as a comparison with a subgroup analysis showed no differences in beta estimates, standard errors, or p-values. Third, a dichotomized outcome of CKD defined as eGFR <60 ml/min/1.73m2 was used. Pattaro *et al.* had a sample size of 117,165, including 12,385 CKD cases from 49 studies, all of European ancestry [29]. Furthermore, Wuttke *et al.* reported GWAS data based on a meta-analysis of over 1 million individuals from the CKDGen Consortium, comprising 41,395 cases and 439,303 controls of European descent [30]. These datasets provided robust and complementary evidence on genetic associations with CKD. A summary of the outcome datasets used in this study is provided in Table 2.

### Mendelian Randomization Analyses

We conducted univariable and multivariable MR analyses to estimate the causal effects of BMI, as well as brain-and adipose-tissue-instrumented BMI, on kidney function outcomes. For univariable MR, we firstly estimated the total effect of genetically predicted adult BMI using the full set of 915 instruments (i.e., without considering their tissue-dependent effects on gene expression) on the kidney outcomes which have previously been shown to be influenced by adiposity, i.e. CKD defined as eGFR <60 ml/min/1.73m2. We used inverse-variance weighting (IVW) as the primary method. We then utilized adipose-and brain-specific expression variants, colocalized with BMI (PPA4 ≥ 0.8), as instrumental variables in the MVMR framework. We termed BMI based on adipose variants as “adipose-tissue instrumented BMI,” and BMI based on brain variants as “brain-tissue instrumented BMI.”

Univariable MR was then conducted to estimate their separate effects on kidney outcomes. Using MVMR, we simultaneously modelled tissue-specific genetic instruments to disentangle the independent effects of brain-and adipose-tissue-related BMI. This was achieved by including the full set of adipose and brain tissue derived instruments for BMI in the MVMR model, using the differential evidence for colocalization between tissues to differentially weight each instrument’s effect size in each tissue-partitioned exposure respectively [14,19]. In both univariable and multivariable analyses, causal effects were estimated using the inverse variance weighted (IVW) method. The IVW approach combines the individual associations of each genetic variant by dividing its gene-outcome association by its gene-exposure association, with each ratio weighted by the inverse of its variance. When all genetic variants are valid instruments, the IVW estimate is the most statistically powerful method. In MVMR, the assumptions are: (i) the SNPs strongly predict each exposure conditional on the others (MV-IV1); (ii) the SNPs are independent of the outcome once all exposures are included (MV-IV2); and (iii) the SNPs are independent of confounders of any exposure–outcome relationship (MV-IV3) [31]. We evaluated instrument strength using conditional F-statistics (MV-IV1). Assumptions MV-IV2 and MV-IV3 are more difficult to verify directly; we sought to address them by including correlated exposures jointly in the model to account for potential pleiotropic pathways, nevertheless formal tests for residual pleiotropy can be performed [15,31,32].

Risk alleles for brain-and adipose-tissue instrumented BMI were harmonized with those for the outcomes of each kidney measure. A harmonized dataset containing consistent reference and alternative alleles was then analyzed. The conditional F-statistic was calculated to assess the strength of genetic instruments for brain and adipose tissue [33]. Valid instruments provide independent and unbiased causal estimates of the relationship between exposure and outcome. The conditional F-statistic for adipose tissue, conditioned on brain tissue, was greater than 10, indicating that bias due to weak instruments was unlikely (Supplementary Table 1).

### Sensitivity Analyses

To evaluate the robustness of IVW results in both univariable and MVMR analyses, sensitivity analyses were conducted. For univariable MR, these included calculating MR-Egger estimates to assess potential directional pleiotropy [34], weighted mode and weighted median methods were applied to further explore horizontal pleiotropy (Supplementary Table ST3), where genetic variants associated with the exposure of interest may influence the outcome through pathways unrelated to the exposure. Global heterogeneity among instrumental variables was assessed using Cochran’s Q statistic. A Q value exceeding L−1(where L represents the total number of instrumental variables) signifies the presence of heterogeneity among the instrumental variables included in the analysis [35]. All analyses were performed using the R packages “TwoSampleMR 0.6.15” [36] and “MendelianRandomization” [37] under R version 4.4.1.

We performed a further analysis investigating the effects of BMI associated genetic instruments stratified by pathway (based on Mendelian disease categories using MendelVar) [38,39]. The pathway-stratified MR approach annotates genetic instruments for complex traits to pathways informed both by the enrichment of Mendelian disease genes near GWAS loci and the alignment of genetic associations to shared phenotypes or symptoms of monogenic and complex forms of disease [40,41,42]. Comparison with this alternative instrument stratification method was to determine whether these external ontologies might offer orthogonal evidence to help dissect the association between BMI and the selected kidney function outcomes. For BMI the most significantly enriched categories were “disease of metabolism” (45 SNPs) and “disease of mental health”/“developmental disorder of mental health” (39 SNPs) [38,43]. This ontology selection enabled us to assess within an MR framework whether the overall effect of BMI on kidney function may be differentially influenced via genetic variants significantly enriched within mental health or metabolic pathways. Notably, of the 45 “mental health” pathway SNPs, only 5 overlapped with tissue-assigned SNPs from Leyden *et al.* [14]—4 with “brain” and 1 with both “adipose” and “brain.” In contrast, 17 of the 39 “metabolic” pathway SNPs showed overlap, including 3 with “adipose,” 5 with both tissues, and 9 with “brain.” This overlap between pathway-stratified “metabolic” SNPs and tissue-stratified “brain” SNPs compared to those associated with “mental health” reflects the fact that the brain eQTL reference panel emphasized regions governing energy balance and homeostatic control [21].

## Results

Univariable IVW MR provided strong evidence of an effect of BMI (number SNPs=915) on CKD as expected 9(OR = 1.24, 95% CI: 1.2 to 1.3) (Table S1). A similar association was observed for brain-tissue instrumented BMI (ORwuttke = 1.25, 95% CI: 1.02 to 1.54, ORpattaro = 1.29, 95% CI: 1.1 to 1.513, respectively) [33, 34] and for adipose-instrumented BMI (ORwuttke = 1.31, 95% CI = 1.06 to 1.62) (Table S1, Figure 1A). Overall BMI and brain-and adipose-instrumented pathways generally were associated with worse kidney function. The association between BMI and eGFRcys was notably stronger and more precise than for eGFRcrea. BMI was inversely associated with eGFRcys (beta = –0.05, 95% CI: – 0.06 to –0.046) (Table S1), and this effect persisted for both brain-instrumented BMI (beta= - 0.05, 95%CI =-0.06 to-0.04) and adipose-instrumented BMI (beta=-0.05, 95% CI=-0.07 to - 0.3) (Table S1). In contrast, no clear associations were observed with eGFRcrea for either tissue-partitioned exposure (Table S1). Brain-and adipose-instrumented BMI also showed a similar association with slower annual decline in eGFR (beta_brain_ = 0.07, 95% CI: 0.01 to 0.13, beta_adipose_ =0.08, 95% CI= −0.007 to 0.17) (Table S1, Figure 1B).

**Figure 1.**
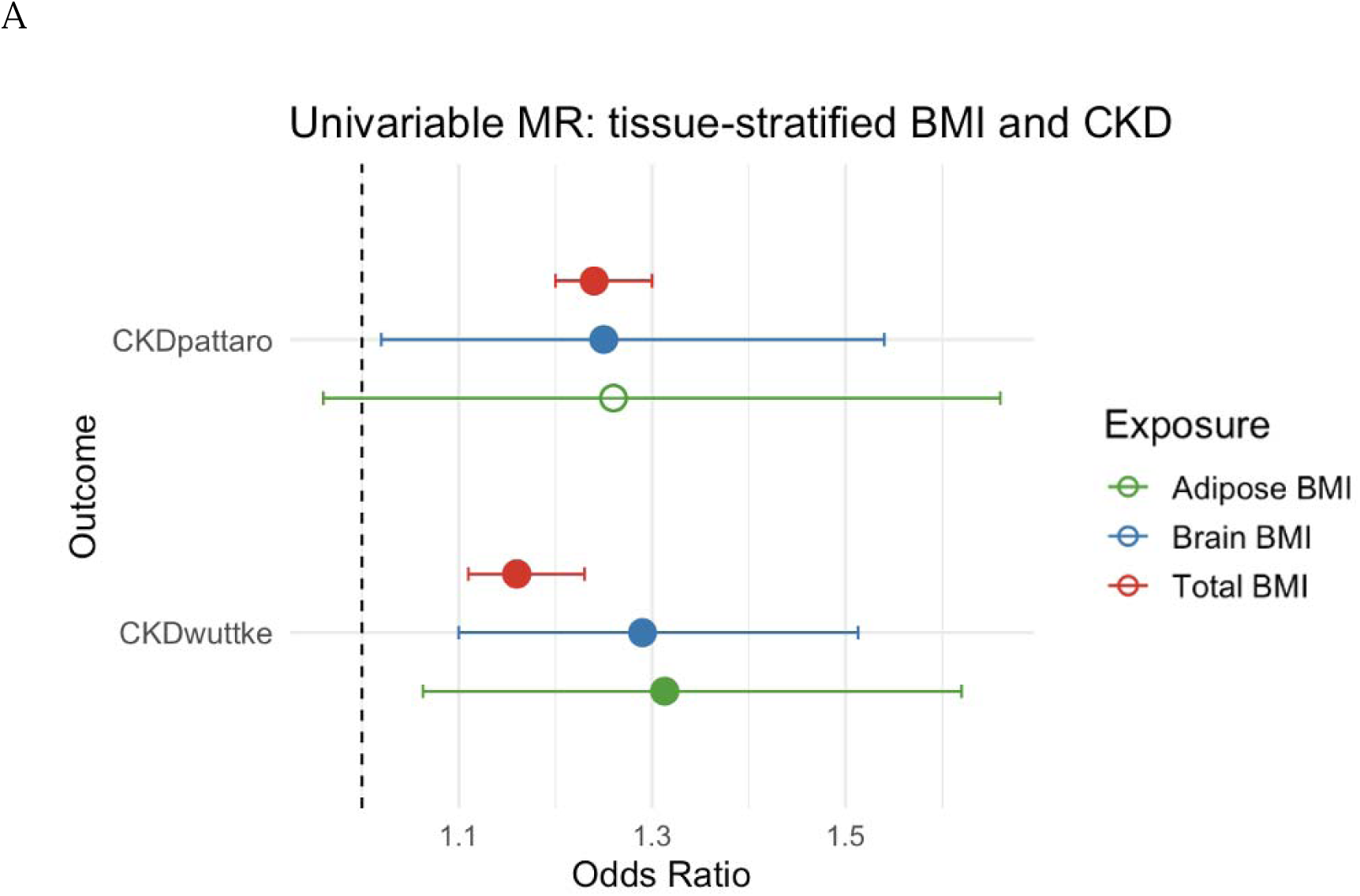
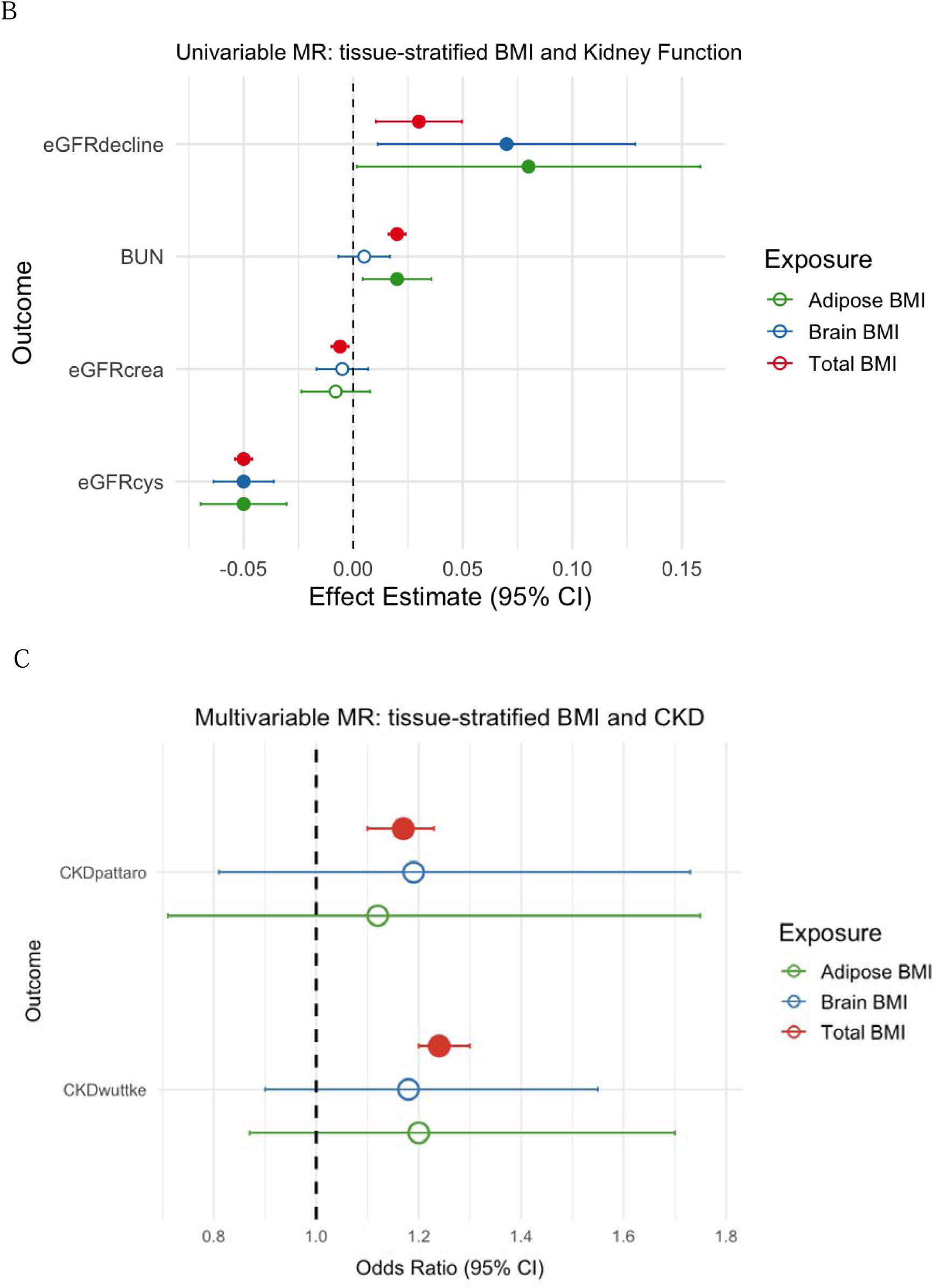
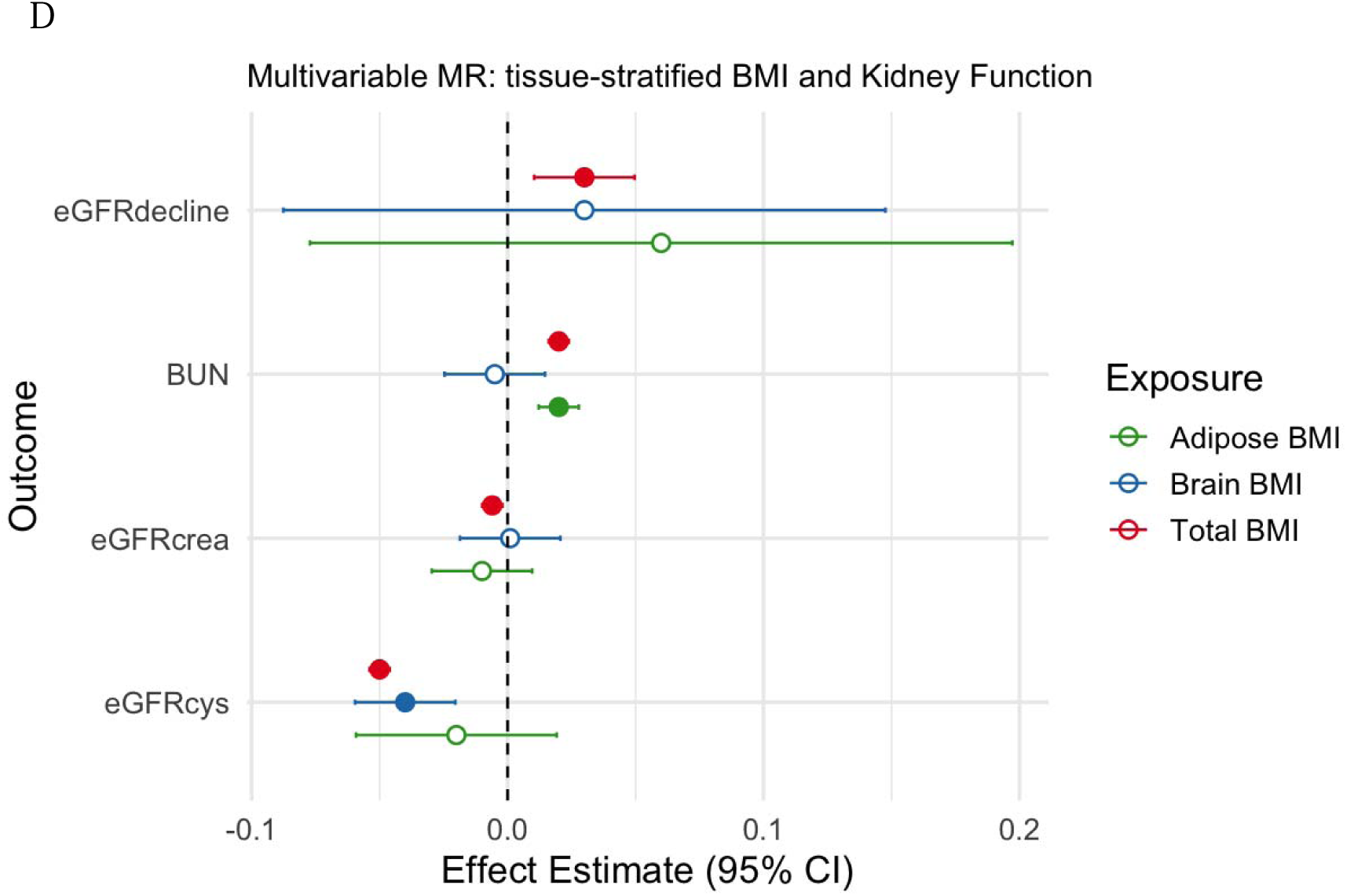
Forest plots of total, adipose-and brain-instrument BMI on CKD and kidney function

In MVMR, after mutually adjusting for either brain or adipose, results were in a concurrent direction with the univariate analysis, with neither showing a stronger effect than the other (e.g, for CKD (Pattaro *et al*.) OR_brain =1.19 [95% CI: 0.81 to 1.73] and OR_adipose = 1.12 [95% CI: 0.71 to 1.75]) (Table S2, Figures 1C, 1D).

Conditional F-statistics for both exposures were above 10, indicating no weak instrument bias [Table S3]. Cochran’s Q statistics from all analyses was p < 2.6 × 10LJ2 (Table S4) suggesting heterogeneity among the complete set of 915 BMI-associated SNPs as well as within the brain and adipose instruments across all examined outcomes.

Forest plots of Mendelian randomization (MR) results for BMI on 2 CKD outcomes (studies) and 4 kidney outcomes using (A and B) univariable analyses using the total set of BMI variants as well as brain and adipose variants (C and D) analyses instrumented in a multivariable setting with tissue-partitioned variants along with effect estimates of BMI instrumented with all 915 BMI SNPs for comparison. Odds ratios or effect estimates per standard deviation (SD) change in exposure and 95% confidence intervals (CIs) for each disease outcome analyzed by MR are shown. BMI, body mas index; CKD, Chronic Kidney Disease; BUN, blood urea nitrogen; eGFRcrea, creatinine-based eGFR; eGFRcys, cystatin based eGFR; 95% CI, 95% confidence interval.

### No Evidence for Independent Pathways

We further examined whether pathway-stratified BMI instruments enriched for variants aligning to “mental health” and “metabolism” related pathways were independently related to kidney function traits [38]. In univariable MR, the “mental health”-related instrument set showed associations with both CKD (Wuttke) (OR = 1.50, 95% CI: 1.15 to 1.85) and eGFRcys (beta = –0.05, 95% CI: –0.07 to –0.040) (Table S4). However, no clear associations were observed for BUN, eGFRcrea, or eGFR decline (Table S4). The “metabolism” pathway also displayed a modest effect on eGFRcys (beta = –0.03, 95%CI = 0.05 to-0.01), but had no clear associations with other outcomes (Table S4). These findings are directionally consistent with our primary brain-and adipose-partitioned univariable and MVMR results, which also similarly demonstrated no clear dominance of one tissue pathway over the other. When both exposures were modelled simultaneously in MVMR, all associations attenuated toward the null (Table S5). Crucially, neither pathway ontology-based exposure exhibited a dominant or independent effect across kidney outcomes in the MVMR framework.

## Discussion

### Summary

Overall BMI was associated with kidney function in univariable analysis, and notably the brain-and adipose-partitioned instruments produced consistent effect sizes, suggesting comparable contributions from both pathways. Our study suggested no clear distinction between adipose and brain tissue mechanisms being predominantly responsible for the underlying pathway between BMI and measures of kidney function. The attenuation in the MVMR results compared to the univariable results suggests that while higher BMI is causally linked to kidney dysfunction, the specific contributions of brain-and adipose-mediated pathways may contribute jointly and their independent effects could not be statistically distinguished. Notably, only brain-instrumented BMI exhibited a consistent association with lower eGFRcys across both univariable and multivariable models. In contrast, its associations with eGFRcrea, BUN, and eGFR decline closely mirrored those of adipose-instrumented BMI, offering no clear indication of a dominant tissue mediated effect. The consistent associations for brain-instrumented BMI with eGFRcys could be compatible with central mechanisms, such as neuroendocrine regulation of appetite.

Our univariable MR analyses leveraging gene ontology–based instruments for “disease of mental health” and “disease of metabolism” pathways revealed directionally consistent associations with kidney function markers, particularly eGFRcys. Specifically, “disease of mental health”—serving as a pathway-stratified BMI instrument enriched for variants aligning to “mental health” —demonstrated a negative effect on eGFRcys, mirroring the inverse relationship observed in our primary brain-tissue partitioned analyses. Similarly, “disease of metabolism”, reflective of pathway-stratified BMI instruments enriched for variants aligning to metabolism, showed effects in the same direction, although modest.

These findings provide corroboration for the tissue-partitioned results, reinforcing the hypothesis that both adipose-and brain-instrumented BMI pathways act as component causes on kidney function.

### Comparison with previous literature

No previous studies have investigated the tissue partitioned effects of BMI on kidney outcomes. The available literature shows that the causal relationship between BMI and CKD is largely mediated by overall adiposity and its metabolic sequelae [44, 45, 46]. However, unlike prior research demonstrating differential associations between tissue-partitioned BMI and cardiometabolic or cancer outcomes [14, 19], our findings did not support distinct causal roles for these BMI components in kidney dysfunction. Nevertheless, the current study differs from previous research by directly testing the causal roles of brain-and adipose-tissue-derived BMI in kidney dysfunction, rather than relying on phenotypic or depot-specific measures. Studies partitioning BMI into fat mass and fat-free mass, or examining specific adipose depots, have consistently shown that visceral adiposity is more strongly associated with reduced GFR than subcutaneous adiposity or overall BMI [45, 46]. Mendelian randomization analyses further support a causal relationship between visceral adipose tissue (VAT) and CKD, with hypertension mediating a substantial proportion of this effect [47].

Furthermore, studies describe a bidirectional brain-adipose-kidney axis, where signals from adipose tissue can modulate renal sympathetic nerve activity via sensory neurons, and the brain can influence renal function through autonomic pathways [48]. This neuroendocrine crosstalk is implicated in the pathogenesis of obesity-related kidney dysfunction.

Additionally, the kidney itself can signal to the brain to regulate feeding and body weight, for example via growth differentiation factor 15, highlighting the complexity of these interactions [49].

### Potential mechanisms

A key factor contributing to the absence of dominance of any of the brain and adipose pathways may be the integrative nature of the kidney as a target organ with multiple upstream pathways converging to affect renal physiology, complicating efforts to isolate single-tissue drivers. Unlike metabolic or cardiovascular outcomes—such as type 2 diabetes or coronary artery disease—which often reflect dominant contributions from discrete physiological systems (e.g., insulin signaling or lipid metabolism), kidney function is influenced by a wide array of systemic processes. These include hemodynamic stress, oxidative damage, inflammatory cascades, endothelial dysfunction, and hormonal dysregulation, all of which can act in concert or in succession to compromise renal function. Consequently, the effect of BMI on CKD may not be readily attributable to a singular tissue of origin but instead reflects a summative burden that is more effectively captured by total adiposity measures [50].

Beyond brain and adipose tissues, other pathways may be pivotal in mediating the impact of BMI on renal health. Hepatic steatosis and impaired liver function, which are common sequelae of obesity, are implicated in the propagation of systemic inflammation and insulin resistance, both of which adversely affect renal microcirculation [51, 52]. Additionally, the role of skeletal muscle mass and composition (i.e., sarcopenic obesity), gut-derived uremic toxins, and vascular endothelial integrity warrant further exploration. Dysregulation of endocrine systems such as the renin–angiotensin–aldosterone system (RAAS) and aberrant leptin signaling may also serve as intermediary mechanisms linking excess adiposity to glomerular hyperfiltration and subsequent nephron damage.

It is also important to acknowledge that the adipose tissue instruments employed in this study were derived from subcutaneous fat depots. As such, the findings may not be generalizable to other adipose compartments, particularly visceral adipose tissue, which differs in metabolic activity, anatomical location, and its potential role in mediating renal risk [53].

Finally, the gene-environment equivalence assumption [54] in MR suggests that the impact of BMI on disease risk may be consistent across different genetic pathways. However, null results in MVMR do not necessarily rule out underlying biological specificity, as instrument strength or overlap between the genetic instruments may obscure distinct causal contributions. The methods applied here exemplify how biological and genetic data can help disentangle the complex architecture of composite traits like BMI. More refined biological insights may emerge from integrating multi-omics and tissue-or cell-type-specific data, which could help isolate more specific mechanisms linking adiposity to kidney function.

### Strengths and Limitations

A key strength of this study is its novel approach—the first of its kind to investigate the effects of tissue-partitioned BMI on CKD using MR. This study leverages large-scale, high-quality genomic data from some of the most comprehensive and well-powered datasets. By integrating these resources, we ensure both statistical power and biological specificity, which enhances the interpretability and translational potential of our findings. Nevertheless, our analyses were primarily based on European-ancestry populations, which may limit the generalizability of these findings to other ancestral groups.

While adipose and brain tissues were the primary focus of this study due to their biological relevance to BMI and the availability of sufficiently powered datasets, they may not necessarily represent the principal tissues through which our colocalized variant sets exert their effects. Although our MR framework leverages the tissue specificity of these instruments, a more comprehensive characterization of each BMI-associated SNP necessitates further investigation utilizing datasets from a broader range of tissues. Within the adipose-and brain-tissue-specific instrument subsets, we also observed considerable heterogeneity as indicated by Cochran’s Q statistics, likely reflecting locus-specific pleiotropy or multi-tissue expression, complicating interpretation of single-tissue pathways. MVMR reduces bias from pleiotropy via measured exposures as it includes all relevant exposures in the model, so the estimated effect of each exposure is conditional on the others, but does not completely remove pleiotropy, especially from unmeasured pathways.

Sensitivity analyses for pleiotropy in MVMR are currently limited; however, multivariable MR-Egger [53] can be applied to account for directional pleiotropy that is uncorrelated with SNP–exposure associations.

Additionally, in patients with established CKD, especially in advanced stages (eGFR <30 mL/min/1.73 m²), the association between high BMI and adverse renal outcomes is attenuated or even absent, a phenomenon sometimes referred to as the “obesity paradox” [55–57]. Our analysis included kidney function measurements in the general population that may not fully reflect disease progression.

One potential source of systematic bias in our analysis arises from the use of eGFRcrea as a measure of kidney function. Creatinine levels are influenced not only by renal function but also by muscle mass. This introduces the possibility of information bias or confounding, whereby part of the observed association between BMI and eGFRcrea may reflect variation in muscle mass rather than true differences in kidney function. Our results suggested that higher BMI—total or tissue-partitioned—negatively impacts kidney function as measured by cystatin C. While eGFRcrea may be biased due to its dependence on muscle mass, eGFRcys offers a more accurate and muscle-mass-independent assessment of kidney function in the context of BMI [58].

A limitation of many CKD GWASs, including those used here, is that CKD is often defined solely based on eGFR < 60 ml/min/1.73m². While this lab-based threshold allows for broader case identification, it may include individuals without clinically confirmed disease. In contrast, population-based studies with registry linkage, such as HUNT [59], enable case definitions based on ICD-coded diagnoses, offering a stricter and more specific classification of CKD.

Finally, exploring sex-specific patterns of adiposity represents a critical direction for future investigation, given the well-established biological differences in fat distribution between males and females [60]. The present study employed non-sex-stratified analyses. Looking ahead, leveraging sex-specific eGFR and CKD data from resources such as the HUNT study [59] could provide valuable insight into whether sex-specific BMI and its tissue-specific components differentially impact renal function across sexes. However, the limited public accessibility of tissue-specific eQTL datasets remains a significant barrier to fully realizing such sex-stratified analyses [61].

### Future Prospects

Clinically, these findings reinforce that BMI reduction remains an important strategy for preserving kidney function, despite not indicating targeting specific brain-or adipose-mediated mechanisms. The lack of distinct effects in MVMR might suggest that future therapeutic strategies should address BMI holistically rather than focusing on tissue-specific mechanisms when considering kidney disease prevention. However, effects in advanced CKD may differ, and interventions should be individualized. Future research should develop more biologically specific instruments to better target pathways linking BMI to renal dysfunction and tailor prevention and treatment strategies accordingly.

## Data Availability

All summary-level data analysed in this study is publicly available.

## Author contributions

AB, HR, and BB conceived and design the study. AB analysed the data. AB wrote the first draft of the manuscript. All authors contributed to the interpretation and revision of the manuscript.

## Ethical standards

We used publicly available summary statistics for this study.

## Data Availability

All summary-level data analysed in this study is publicly available.

## Acknowledgements

We thank the genome-wide association study consortia who made their summary statistics publicly available for this study. We thank the team in the Chronic Kidney Disease Genetics Consortium and Medical Research Council Integrative Epidemiology Unit at the University of Bristol for access to the GWAS summary statistics used in this study.

