## Supplementary Tables for "Exploring the Causal Relationship Between Body Mass Index and Kidney Function Using Tissue-Partitioned Mendelian Randomization"

Table S1. Univariable Inverse Variance Weighted (IVW) estimates of “brain- “and “adipose- tissue instrumented BMI” on Kidney Function markers

| Outcomes | Exposures | SNPs | Beta/OR* | Se | 95% CI | P-value |
| --- | --- | --- | --- | --- | --- | --- |
| CKD  (eGFRcrea <60 ml min^−1^ per 1.73 m^2^) | BMI | 881 | 1.24 | 0.04 | 1.2 to 1.3 | 9 x 10^-8^ |
|  | BMI_brain | 140 | 1.25 |  | 1.02 to 1.54 | 0.03 |
|  | BMI_adipose | 88 | 1.26 |  | 0.96 to 1.66 | 0.09 |
| eGFR(decline) | BMI | 891 | 0.03 | 0.01 | 0.005 to 0.049 | 0.3 |
|  | BMI_brain | 140 | 0.07 | 0.03 | 0.01 to 0.13 | 0.02 |
|  | BMI_adipose | 87 | 0.08 | 0.04 | −0.007 to 0.17 | 0.07 |
| eGFRcys | BMI | 888 | -0.05 | 0.002 | -0.06  to -0.046 | 10^-115^ |
|  | BMI_brain | 140 | -0.05 | 0.007 | -0.06 to -0.04 | 2 x 10^-13^ |
|  | BMI_adipose | 87 | -0.05 | 0.01 | -0.07 to -0.3 | 3,4 x 10^-5^ |
| eGFRcrea | BMI | 890 | -0.006 | 0.002 | -0.009  to -0.002 | 0.001 |
|  | BMI_brain | 140 | -0.005 | 0.006 | -0.02 to 0.006 | 0.35 |
|  | BMI_adipose | 87 | -0.008 | 0.008 | -0.02 to 0.008 | 0.33 |
| BUN | BMI | 886 | 0.02 | 0.002 | 0.01  to 0.021 | 6.4 x 10^-13^ |
|  | BMI_brain | 140 | 0.005 | 0.006 | -0.006 to 0.02 | 0.4 |
|  | BMI_adipose | 87 | 0.02 | 0.008 | 0.002 to 0.03 | 2.5 × 10⁻² |
| CKD (Wuttke) | BMI | 889 | 1.16 | 0.03 | 1.1 to 1.23 | 1.8 x 10 ^-07^ |
|  | BMI_brain | 140 | 1.29 |  | 1.1 to 1.513 | 0.002 |
|  | BMI_adipose | 87 | 1.313 |  | 1.063 to 1.62 | 0.01 |

* The causal estimates are presented as beta with 95% CI for all outcomes besides CKD, which is presented as odds ratio with 95% CI.

**Table S2. Multivariable causal estimates of “brain- “and “adipose- tissue instrumented BMI” on Kidney Function markers in a two-sample setting (PPA>0.8)**

| **Outcomes** | **Exposures** | **SNPs** | **Beta/OR*** | **Se** | **95% CI** | **P-value** |
| --- | --- | --- | --- | --- | --- | --- |
| **CKD (Pattaro *et al.)***  **(eGFRcrea <60 ml min^−1^ per 1.73 m^2^)** | BMI_brain | 182 | 1.19 | 0.23 | 0.81 to 1.73 | 0.36 |
|  | BMI_adipose | 182 | 1.12 | 0.19 | 0.71 to 1.75 | 0.61 |
| **eGFR(decline)** | BMI_brain | 183 | 0.03 | 0.06 | -0.08 to  0.14 | 0.55 |
|  | BMI_adipose | 183 | 0.06 | 0.07 | -0.06 to 0.19 | 0.33 |
| **eGFRcys** | BMI_brain | 182 | -0.04 | 0.01 | -0.07 to -0.01 | 0.003 |
|  | BMI_adipose | 182 | -0.02 | 0.02 | -0.06 to 0.01 | 0.18 |
| **eGFRcrea** | BMI_brain | 182 | 0.001 | 0.01 | -0.02 to 0.02 | 0.91 |
|  | BMI_adipose | 182 | -0.01 | 0.01 | -0.04 to 0.01 | 0.27 |
| **BUN** | BMI_brain | 182 | -0.005 | 0.01 | -0.03 to 0.01 | 0.58 |
|  | BMI_adipose | 182 | 0.02 | 0.01 | -0.004 to 0.04 | 0.096 |
| **CKD (Wuttke *et. al)*** | BMI_brain | 183 | 1.18 | 0.14 | 0.9 to 1.55 | 0.23 |
|  | BMI_adipose | 183 | 1.2 | 0.2 | 0.87 to 1.7 | 0.26 |

* The causal estimates are presented as beta with 95% CI for all outcomes besides CKD, which is presented as odds ratio with 95% CI.

Table S3: Conditional F-statistics of the genetic instruments

| **Outcome** | **Exposure** | **Multivariable MR** |
| --- | --- | --- |
| CKD (Wuttke *et al.)* | BMI_brain | 35.90882 |
|  | BMI_adipose | 26.32063 |
| eGFRcys | BMI_brain | 35.90882 |
|  | BMI_adipose | 26.32063 |
| eGFRcrea | BMI_brain | 35.90882 |
|  | BMI_adipose | 26.32063 |
| BUN | BMI_brain | 35.90882 |
|  | BMI_adipose | 26.32063 |
| eGFR(decline) | BMI_brain | 35.87168 |
|  | BMI_adipose | 26.75384 |

Table S4. Heterogeneity test of univariable MR

| **Outcome** | **Exposure** | **Q (P-value)** |
| --- | --- | --- |
| CKD (Wuttke *et al.)* | BMI_brain | 139 (1.1x10^-8^) |
|  | BMI_adipose | 173 (7.5x10^-8^) |
|  | BMI | 1323 (1.7x10^-13^) |
| eGFRcys | BMI_brain | 144 (9x10^-14^) |
|  | BMI_adipose | 406 (2x10^-74^) |
|  | BMI | 1141 (6x10^-111^) |
| eGFRcrea | BMI_brain | 1342 (5.8x10^-196^) |
|  | BMI_adipose | 997 (4.5x10^-155^) |
|  | BMI | 5133 (0) |
| BUN | BMI_brain | 469 2x10^-37^) |
|  | BMI_adipose | 337 (1.9x10^31^) |
|  | BMI | 3977 (0) |
| eGFR(decline) | BMI_brain | 139 (3x10^-4^) |
|  | BMI_adipose | 139 (2x10^-4^) |
|  | BMI | 1058 (0.02) |
| CKD (Pattaro *et al.)* | BMI_brain | 139 (0.1) |
|  | BMI_adipose | 110 (0.04) |
|  | BMI | 967 (0.3) |

Table S5 Univariable Inverse Variance Weighted (IVW) estimates of “disease of mental health “and “disease of metabolism” gene ontologies on Kidney Function markers

| **Outcomes** | **Exposures** | **SNPs** | **Beta/OR*** | **Se** | **95% CI** | **P-value** |
| --- | --- | --- | --- | --- | --- | --- |
| BUN | “disease of mental health”    “disease of metabolism” | 43 | 0.0027 | 0.01 | -0.02 to 0.0028 | 0.8 |
|  |  | 37 | -0.03 | 0.01 | -0.05 to -0.01 | 0.07 |
| CKD Wuttke | “disease of mental health” | 43 | 1.5 | 0.12 | 1.15 to 1.85 | 0.002 |
|  | “disease of metabolism” | 37 | 1.1 | 0.16 | 0.88 to 1.38 | 0.4 |
| eGFRcys | “disease of mental health” | 43 | -0.05 | 0.007 | -0.07 to -0.04 | 1,6 x 10-12 |
|  | “disease of metabolism” | 37 | -0.03 | 0.01 | -0.05 to -0.01 | 0.009 |
| eGFRcrea | “disease of mental health” | 43 | -0.01 | 0.008 | -0.03 to 0.005 | 0.2 |
|  | “disease of metabolism” | 37 | 0.006 | 0.007 | -0.009 to 0.02 | 0.44 |
| eGFR decline | “disease of mental health” | 43 | 0.03 | 0.06 | -0.08 to 0.15 | 0.6 |
|  | “disease of metabolism” | 37 | 0.06 | 0.06 | -0.05 to 0.16 | 0.3 |
| CKD Pattaro | “disease of mental health” | 43 | 1.4 | 0.19 | 0.97 to 2.04 | 0.07 |
|  | “disease of metabolism” | 37 | 1.2 | 0.18 | 0.87 to 1.7 | 0.28 |

Table S6. Multivariable Inverse Variance Weighted (IVW) estimates of “disease of mental health “and “disease of metabolism” gene ontologies on Kidney Function markers

| **Outcomes** | **Exposures** | **SNPs** | **Beta/OR*** | **Se** | **95% CI** | **P-value** |
| --- | --- | --- | --- | --- | --- | --- |
| **CKD (Pattaro)** | “disease of mental health” | 75 | **0.88** |  | 0.6 to 1.3 | 0.5 |
|  | “disease of metabolism” | 75 | **1.1** |  | 0.8 to 1.6 | 0.6 |
| **eGFR(decline)** | “disease of mental health” | 75 | -0.11 | 0.05 | -0.22 to  -0.01 | 0.03 |
|  | “disease of metabolism” | 75 | -0.03 | 0.05 | -0.13 to 0.07 | 0.53 |
| **eGFRcys** | “disease of mental health” | 75 | -0.009 | 0.01 | -0.03 to 0.01 | 0.4 |
|  | “disease of metabolism” | 75 | -0.010 | 0.01 | -0.03 to 0.01 | 0.4 |
| **eGFRcrea** | “disease of mental health” | 75 | 0.006 | 0.008 | -0.008 to 0.02 | 0.4 |
|  | “disease of metabolism” | 75 | -0.009 | 0.008 | -0.02 to 0.006 | 0.23 |
| **BUN** | “disease of mental health” | 75 | 0.002 | 0.01 | -0.02 to 0.03 | 0.85 |
|  | “disease of metabolism” | 75 | -0.003 | 0.01 | -0.03 to 0.02 | 0.8 |
| **CKD (Wuttke)** | “disease of mental health” | 75 | 1.1 |  | 0.8 to 1.4 | 0.6 |
|  | “disease of metabolism” | 75 | 1.02 |  | 0.8 to 1.3 | 0.9 |

* The causal estimates are presented as beta with 95% CI for all outcomes besides CKD, which is presented as odds ratio with 95% CI.
